# Decomposition of phenotypic heterogeneity in autism reveals distinct and coherent genetic programs

**DOI:** 10.1101/2024.08.15.24312078

**Authors:** Aviya Litman, Natalie Sauerwald, LeeAnne Green Snyder, Jennifer Foss-Feig, Christopher Y. Park, Yun Hao, Ilan Dinstein, Chandra L. Theesfeld, Olga G. Troyanskaya

## Abstract

Unraveling the phenotypic and genetic complexity of autism is extremely challenging yet critical for understanding the biology, inheritance, trajectory, and clinical manifestations of the many forms of the condition. Here, we leveraged broad phenotypic data from a large cohort with matched genetics to characterize classes of autism and their patterns of core, associated, and co-occurring traits, ultimately demonstrating that phenotypic patterns are associated with distinct genetic and molecular programs. We used a generative mixture modeling approach to identify robust, clinically-relevant classes of autism which we validate and replicate in a large independent cohort. We link the phenotypic findings to distinct patterns of *de novo* and inherited variation which emerge from the deconvolution of these genetic signals, and demonstrate that class-specific common variant scores strongly align with clinical outcomes. We further provide insights into the distinct biological pathways and processes disrupted by the sets of mutations in each class. Remarkably, we discover class-specific differences in the developmental timing of genes that are dysregulated, and these temporal patterns correspond to clinical milestone and outcome differences between the classes. These analyses embrace the phenotypic complexity of children with autism, unraveling genetic and molecular programs underlying their heterogeneity and suggesting specific biological dysregulation patterns and mechanistic hypotheses.

## Introduction

At its core, autism spectrum disorder (ASD) is characterized by persistent deficits in social communication and interaction, alongside restricted and repetitive patterns of behavior, interests, or activities^1,2^. The number of ASD diagnoses has been rising rapidly in recent years, and with the widening of diagnostic criteria, there is increasing heterogeneity within the autistic population, phenotypically and genetically^3^. Autism displays a complex phenotypic structure: core features can vary substantially in severity and presentation, and can coincide with an extensive and unique spectra of associated phenotypes and co-occurring conditions for each individual^2,4^. This wide array of phenotypes is matched by the broad genetic heterogeneity of individuals with autism. Despite significant evidence for a genetic basis of the condition^5–18^ and hundreds of ASD-associated genes identified^19–22^, we lack a coherent mapping of genetic variation to phenotypes. Here, we leverage broad phenotypic data on a large autism cohort to parse out both genetic and phenotypic heterogeneity and identify robust phenotypic classes of individuals and their underlying genetic programs.

Previous studies have demonstrated the extensive heterogeneity in autism phenotypes, including in cognitive or adaptive behavior^23,24^, morphology^25^, neuroanatomical imaging profiles^26,27^, and clinical outcomes^28^. However, linking this heterogeneity to genetic factors has been challenging and limited to trait-centric approaches instead of considering the phenotypes of individuals holistically (i.e. the combination of traits of each individual)^12,13,16^. Trait-centric approaches marginalize co-occurring phenotypes when focusing on one trait. However, since traits are not independent, they cannot be separately associated with patterns of genetic variation. Person-centered approaches holistically consider phenotypic traits and their interactions within individuals, and can identify complex genetic patterns that may be associated with these co-occurring phenotypes. This type of approach has shown promise in application to other complex psychiatric conditions^29,30^. During development, traits impact each other in complex ways, compensating for or exacerbating individual phenotype measures. A person-centered approach can capture the sum of these developmental processes at later ages, offering strong clinical value for prognosis with individualized genotype-phenotype relationships. A large sample size with broad phenotyping is necessary to quantitatively analyze phenotypic structures in autism^31,32^. Here, we adopt a person-centered approach to decompose a large sample of individuals and identify reproducible, clinically-relevant classes and their genetic underpinnings.

This quantitative phenotypic analysis enables us to address the long-standing challenge of deconvolving the complexity of genetic signals in autism. We leverage a unique cohort with both broad phenotypic and genotypic data at scale (n = 5,392) in order to parse heterogeneity consistent with clinically meaningful presentations of autism. Using a generative mixture modeling framework, we decompose phenotypic information to identify, validate, and replicate four latent classes, allowing us to associate each of them with distinct and coherent genetic programs. We conduct a thorough investigation of both rare and common genetic influences in the context of the phenotypic heterogeneity defining our classes, showing that patterns in common genetic variation measured by polygenic scores (PGS) coincide with their phenotypic and diagnostic traits. We then analyze *de novo* and rare inherited variation and identify distinct genetic profiles corresponding to clinical outcomes. Finally, we demonstrate that rare variation is associated with class-specific gene expression patterns during development that align with clinical milestones and individual presentations of autism (Figure 1A).

**Figure 1.**
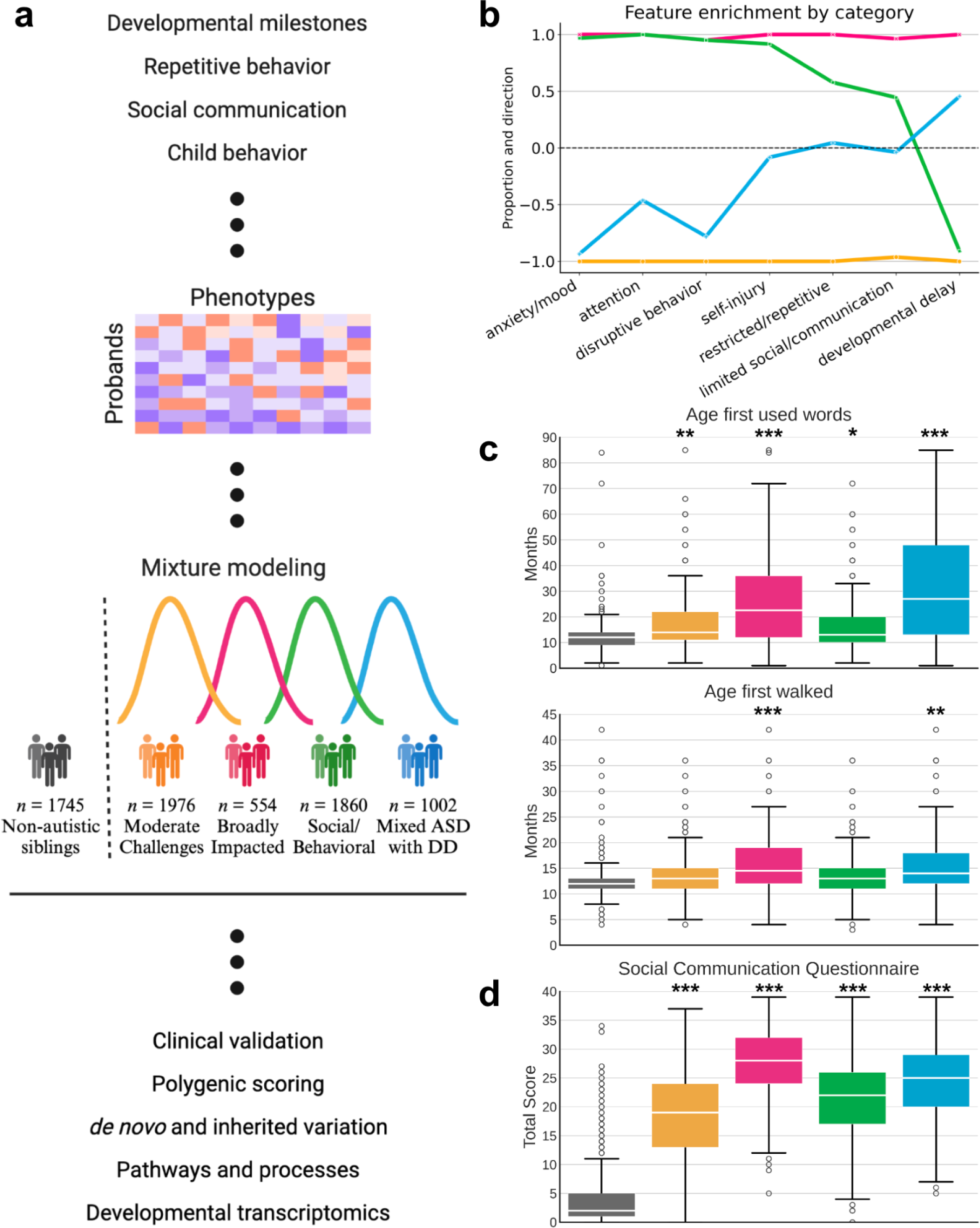
**a,** Study design for parsing the phenotypic heterogeneity of autism and deciphering the genetic factors contributing to individual presentations. A matrix of probands by phenotypes was constructed, with 239 features describing item-level and composite (summary score) phenotypic measure data (developmental milestones, repetitive behavior, social communication, and emotional and behavioral traits) across 5,392 individuals with complete feature data. The 5392 x 239 matrix was used as input to a general finite mixture model to learn the distributions of the latent classes in the input data. We describe four data-driven classes of autism which exhibit distinct phenotypic presentations and trait patterns. We externally validated the classes by showing that their profiles align with clinical data not included in the model training, and performed downstream genetic analyses to associate genetic factors and patterns with each phenotype class. Created with *BioRender*. **b**, To demonstrate differences in phenotype patterns across the four classes, we assess the propensity towards each of 7 phenotype domains with clinical significance for autism. Features were assigned to domains based on published factor analyses of the included questionnaires. For each class, we computed the significance of enrichment in both directions (enriched, depleted) for each feature. Values close to 1 indicate that the majority or all phenotypes within the category are significantly and positively enriched for a given phenotype domain (indicating higher difficulties for that class), and values close to −1 indicate significant negative enrichment or depletion for a given phenotype domain (indicating lower difficulties). The resulting classes are: Broadly Impacted (magenta, n=554), Social/Behavioral (green, n=1860), Mixed ASD with DD (blue, n=1002), and Moderate Challenges (orange, n=1976). **c**, Distributions of two key developmental milestones: age when first walked and age when first used words (both in months) across the four classes, with non-autistic siblings as a control. One-sided independent t-test with multiple hypothesis correction was used to determine significance of enrichment for each class compared to siblings (* indicates FDR < 0.1, ** indicates FDR < 0.05, and *** indicates FDR < 0.01 in all figures). **d**, Individual total scores from the SCQ by class, with non-autistic siblings as a control. The final SCQ score is quantified on a 0-39 scale, with higher scores indicating greater impairment. One-sided independent t-tests with multiple hypothesis correction were used to determine significance of enrichment. Center lines in all boxplots represent the median, box limits represent the upper and lower quartiles, whiskers extend to show the rest of the distribution, and outliers are shown separately.

## Results

### Identifying the structures of autism phenotypes

In order to best reflect the complexity of presentations across autistic individuals, we identified 239 item-level and composite phenotype features present in 5,392 individuals from the SPARK cohort^33^, a nationwide effort to collect and track genetic and clinical presentations over time. Briefly, these features represent responses on standard diagnostic questionnaires: the Social Communication Questionnaire-Lifetime (SCQ)^34^, Repetitive Behavior Scale - Revised (RBS-R)^35^, Child Behavior Checklist 6-18 (CBCL)^36^, and a background history form focused on developmental milestones. These data were analyzed with a General Finite Mixture Model (GFMM) to minimize statistical assumptions while accommodating for heterogeneous (continuous, binary, and categorical) data types (details in Methods). The model aims to capture the underlying distributions in the data and provides an inherently person-centered approach, separating individuals into classes rather than fragmenting each individual into separate phenotypic categories (Figure 1A).

We selected a GFMM with four latent classes representing four distinct patterns of phenotype profiles. After training models with 2-10 latent classes, we found that four classes presented the best balance of model fit as measured by the Bayesian Information Criterion (BIC), Validation Log Likelihood, and other statistical measures of fit (Supplementary Fig. 1; Supplementary Table 1). Additionally, a four class solution offered the best interpretability in terms of phenotypic separation, as evaluated by clinical collaborators with extensive experience working with autistic individuals (Methods). As observed clinically, classes differ not only in severity of autism symptoms, but also in the degree to which co-occurring cognitive, behavioral, and psychiatric concerns factor into their presentation. To highlight the clinical interpretability, we then assigned each of the 239 phenotype features to one of the following seven categories defined in literature^35,37–39^: limited social/communication, restricted/repetitive behavior, attention, disruptive behavior, anxiety/mood, developmental delay, and self-injury (Figure 1B). We identified one class that demonstrated high scores (higher difficulties) across core autism categories of social/communication and restricted/repetitive behaviors, as well as disruptive behaviors, attention, and anxiety, and no reports of developmental delays; this class is referred to as Social/Behavioral (*n* = 1860). A second class shows more nuanced presentation within certain categories, with some features enriched and some depleted among the restricted/repetitive behavior, social/communication, and self-injury categories, and an overall strong enrichment of developmental delays (Figure 1C, Supplementary Fig. 2); this class is called Mixed ASD with DD (*n* = 1002). The last two classes scored consistently lower (fewer difficulties) and consistently higher than other autistic children across all seven categories. These two classes were termed Moderate Challenges and Broadly Impacted, respectively, and represent both the largest (*n* = 1976) and smallest (*n* = 554) of the classes. Note that though the Moderate Challenges class scores below other autistic children across all measured categories, they and all other classes still score significantly higher than non-autistic siblings on the SCQ, the only diagnostic questionnaire with sibling responses, supporting their ASD diagnoses (Figure 1D; Supplementary Table 2).

### Phenotype classes display distinct clinical characteristics and replicate across cohorts

The characteristics of the four phenotypic classes we identified align with data on diagnoses of co-occurring conditions and parent reports that are external to our modeling and class identification. A medical history questionnaire, which includes reports on diagnoses of conditions such as ADHD, OCD, language delays, depression, and anxiety, was not included in the GFMM, but we find that enrichment patterns of these diagnoses match the class-specific phenotypic profiles (Figure 2A). The Broadly Impacted class displays significant enrichment in almost all measured co-occurring conditions, with the Social/Behavioral class only nearing the same diagnostic levels in ADHD, anxiety, and major depression (Figure 2A). These diagnoses reflect the categories with similar phenotype scores between the two classes from the GFMM (Figure 1B). The Mixed ASD with DD class is highly enriched in conditions such as language delay, intellectual disability, and motor disorders, fitting their higher scores in the categories of developmental and restricted/repetitive behaviors, with lower levels of ADHD, anxiety, and depression, as expected based on their phenotypic profile. The two classes with greater developmental delays, Mixed ASD with DD and Broadly Impacted, also show significantly higher parental reports of cognitive impairment, lower levels of language abilities, and much earlier diagnosis ages than the two classes without significant developmental delays (Figure 2B). Furthermore, the average number of interventions such as medication, counseling physical therapy, or other forms of therapy per child is highest among the Broadly Impacted and the Social/Behavioral classes, and lowest in the Moderate Challenges class (Figure 2B). Together, these analyses of medical features show that the four classes identified by the GFMM are phenotypically distinct, supporting their separation in further analyses identifying distinct genetic signals.

**Figure 2.**
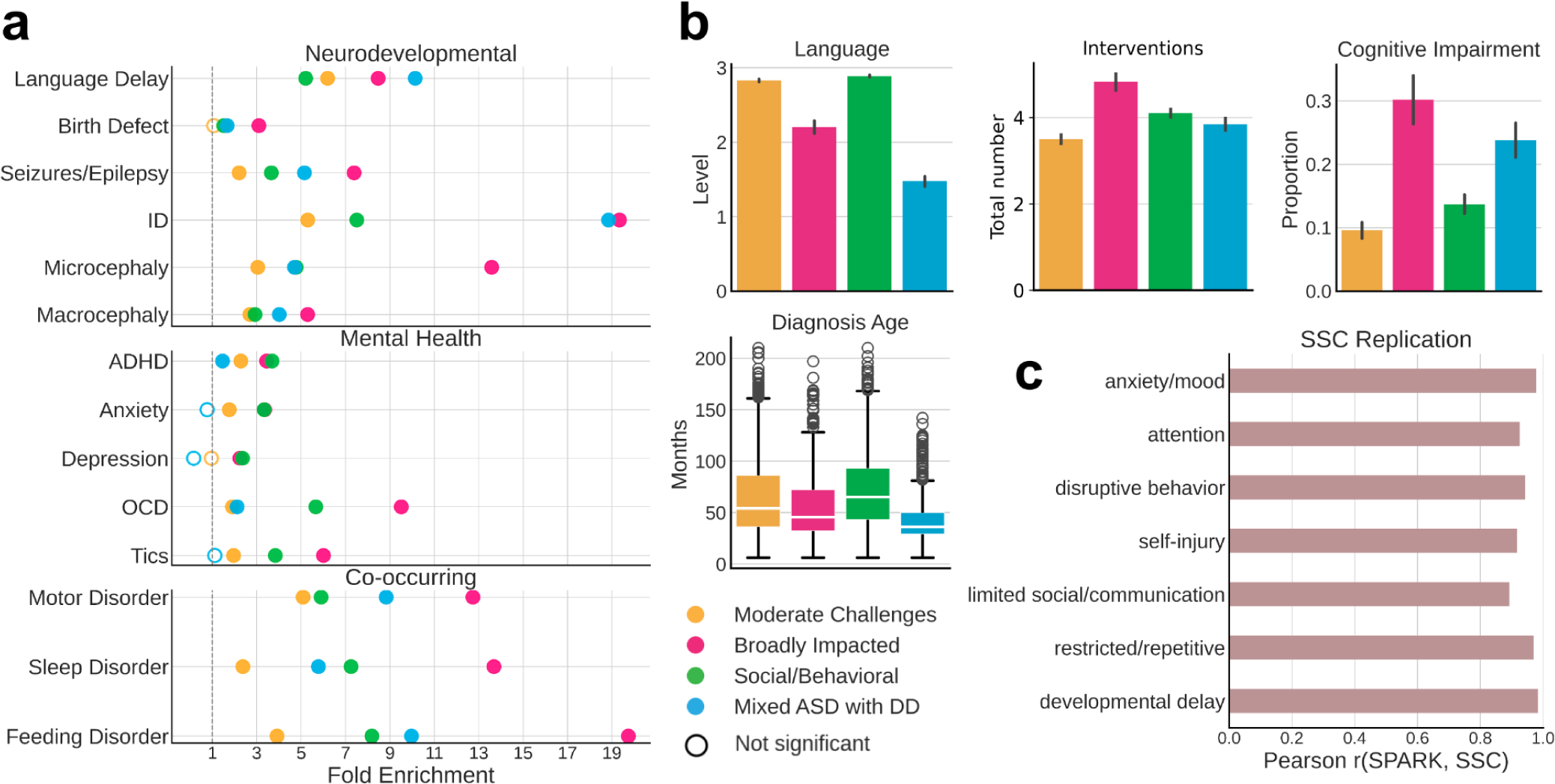
**a**, Clinical validation of classes with external medical diagnoses across three categories: neurodevelopmental, mental health, and co-occurring conditions. We computed the fold enrichment (FE, x-axis) and Benjamini-Hochberg corrected statistical significance (FDR) for a selection of available diagnoses in the Basic Medical Questionnaire for each class. Open circles indicate lack of statistical significance (FDR > 0.05), while closed circles indicate significant enrichments for a diagnosis, and are colored by the corresponding phenotype class. All statistical comparisons were computed with non-autistic siblings as background. The dotted line indicates FE = 1. **b**, External validation of classes with additional parent-reported data from background history and medical history questionnaires. Four variables of interest are displayed: language level at enrollment (parent report with 4 levels reflecting language abilities: 0 = Nonverbal, 1 = Single words, 2 = Phrases, 3 = Sentences), total number of interventions probands in each class have had (including options like medication, physical therapy, social skills groups, speech therapy, recreational therapy, and counseling, among others), cognitive impairment at enrollment (binary indicator of a diagnosis of intellectual disability or cognitive impairment), and diagnosis age in months. Box plot and raw data were plotted for continuous variables, while bar plots showing mean and standard errors were plotted for the binary and categorical variables. **c**, Replication of phenotype classes in an independent cohort – the Simons Simplex Collection. An independent model was trained on the SPARK dataset for 108 features which matched across the measures available for the two cohorts, and was then applied to the SSC dataset. Class labels were obtained for all SSC individuals who had complete data across the 108 features (*n* = 861). Enrichment and depletion of features within each class was computed, and the proportion and direction of enrichment for each of the seven phenotype categories was obtained. The resulting proportions for each category across the 4 classes from SPARK and SSC were correlated (Pearson r, x-axis).

Furthermore, the four phenotype classes replicated well in another independent and deeply phenotyped autism cohort – the Simons Simplex Collection (SSC)^40^, despite a much smaller sample size (*n* = 861). Most phenotypic questionnaires used in the SPARK model were available for SSC, including the SCQ, RBS-R, CBCL (with the exception of item-level data), and a questionnaire on developmental milestones. We combined these matched data, resulting in 108 training features present for both cohorts. To demonstrate the generalizability of our model to the SSC cohort, we applied a GFMM trained on SPARK data to the SSC test set, obtaining class labels for all autistic children in SSC with complete feature data. For quantitative comparisons across cohorts, we computed the enrichment and depletion of each feature within each class across the seven phenotype categories for both cohorts, as described above for the original SPARK model. We demonstrate a strong replication of the autism classes in the SSC cohort, with highly similar feature enrichment patterns across all seven categories (Figure 2C). We further assessed the significance of the overall model similarity with a permutation test, where we randomly shuffled the class labels of the SSC participants 10,000 times, and never observed a higher correlation value with random assignments than the true correlation of 0.927 (p-value < 1e-4, Supplementary Fig. 3). The phenotypic classes defined here are therefore both validated by clinical data and can be replicated in an external cohort.

### Distinct genetic signals underlie phenotypic heterogeneity

We expect that the differences in phenotypes, co-occurring diagnoses, and developmental milestones across the four autism classes correspond to class-specific patterns in genetic signals for common variants. To test this, we computed polygenic scores (PGS) of children with European ancestry for autism and five other well-powered GWAS of related traits and conditions (Supplementary Table 3) indeed, showing significant differences across the four classes that qualitatively match their clinical and phenotypic characteristics (Figure 3A, *n =* 2,293 probands, *n =* 3,179 non-autistic siblings). Interestingly, the class with the highest autism PGS signal is the Moderate Challenges class, demonstrating a differential enrichment of common risk variants in this class compared to the others (FDR = 0.087). The Moderate Challenges class also displays the highest IQ PGS, which makes this finding consistent with previous work demonstrating that ASD PGS positively correlates with intelligence PGS^41^ and coincides with lower likelihood of co-occurring developmental delays^12^. This relatively high signal could also be explained by the GWAS training data most resembling this class phenotypically, while the Broadly Impacted and Mixed ASD with DD classes are poorly represented by the GWAS cohort (Supplementary Fig. 4). The ADHD and Major Depressive Disorder (MDD) PGS signals within the classes matches their diagnostic burden, with Broadly Impacted and Social/Behavioral both showing very high ADHD signal (Broadly Impacted FDR = 2.44e-05, Social/Behavioral FDR = 4.12e-06), and significant enrichment of ADHD diagnoses, while Social/Behavioral shows the highest MDD PGS signal and highest MDD diagnostic burden (Figures 2A and 3A, FDR = 7.14e-4). This demonstrates that co-occurring conditions were associated with common genetic variation that differs across the four identified classes. Additionally, the two classes enriched for intellectual disability and displaying the most significant levels of cognitive impairment (the classes with greater developmental delays) exhibited significantly lower Educational Attainment (Broadly Impacted FDR = 9.78e-08, Mixed ASD with DD FDR = 1.69e-4) and IQ PGS values compared to non-autistic siblings (Broadly Impacted FDR = 1.99e-3, Mixed ASD with DD FDR = 0.079). Only the Moderate Challenges class displayed a depletion in Epilepsy PGS values relative to siblings (FDR = 0.0887), matching the trend demonstrated in previous work in which autistic individuals exhibited lower Epilepsy PGS than non-autistic controls^41^.

**Figure 3.**
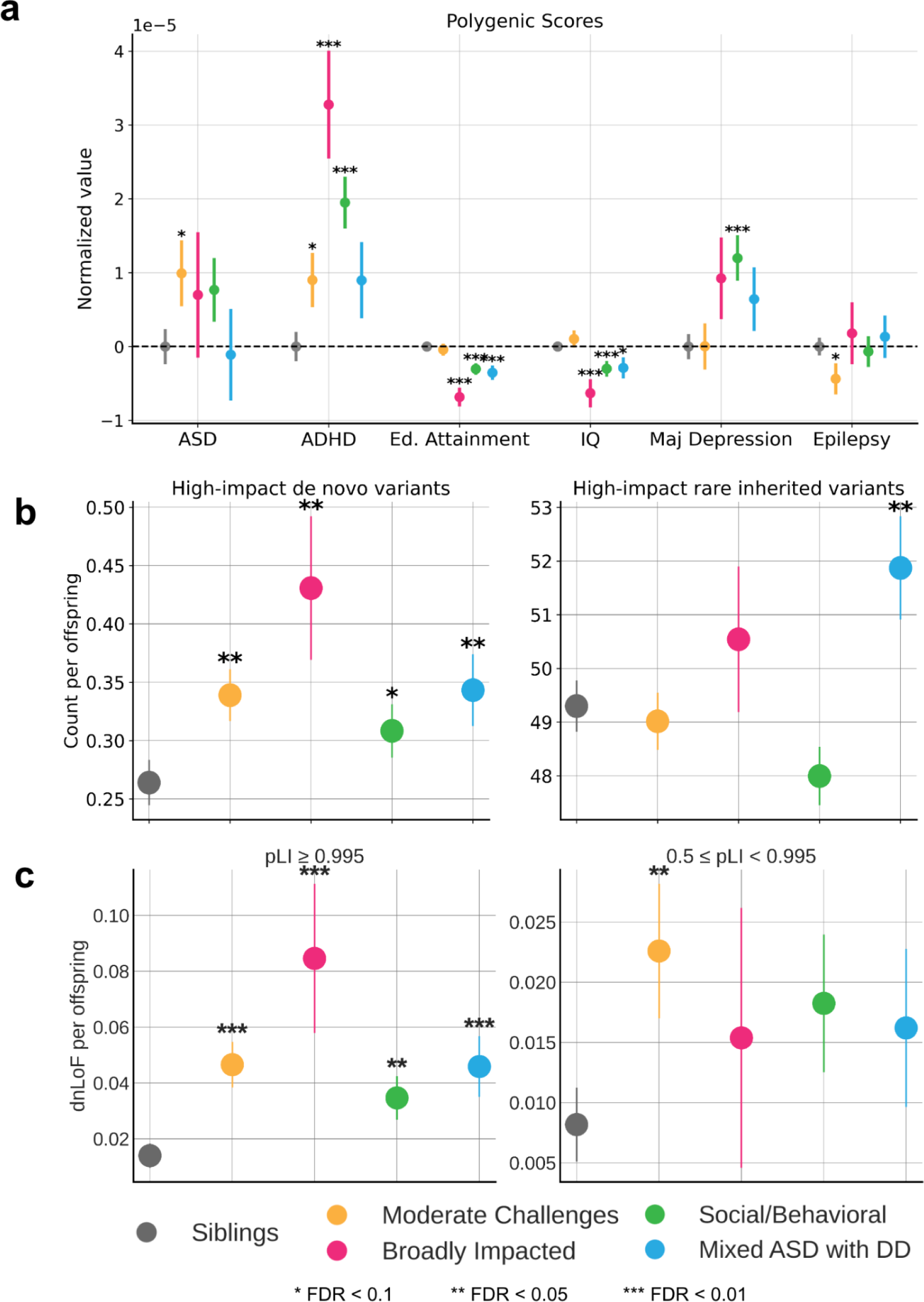
**a**, Polygenic scores (PGS) for ASD GWAS and related phenotypes and conditions. PGS were normalized by the mean of sibling scores within each condition. **b**, Count per offspring of high-impact *de novo* variants (left) and high-impact rare inherited variants (right) across all protein-coding genes. High-impact variants are defined as variants predicted to be either high-confidence loss of function (LoF) or likely pathogenic missense. **c**, Analysis of evolutionarily constrained genes across autism classes and non-autistic siblings. Using the gene-centric measure of evolutionary constraint, pLI, we assigned genes with pLI > 0.5 to one of two classes: pLI > 0.995 (higher constraint genes), or 0.5 <= pLI < 0.995 (lower constraint genes). Count burdens (*de novo* LoF) per offspring were then computed for each class. In all figures, circles indicate the mean and lines show the standard error for each class. Statistical significance is computed with a one-sided independent t-test to compare each class to non-autistic siblings. One star (*) indicates FDR < 0.1, two stars (**) indicate FDR < 0.05, and three stars (***) indicate FDR < 0.01.

In addition to distinct patterns of common variants, we also observed significant differences in rare genetic variation between the phenotypic classes. We conducted *de novo* variant and inherited variant calling on the whole exomes of individuals from the cohort using HAT^42^, and further classified variants as either loss-of-function (LoF), missense, or synonymous. Count-burden enrichments were then computed for each variant type in the four latent classes and compared to non-autistic siblings (details in Methods). We found that the Broadly Impacted class displayed the greatest enrichment for high-confidence *de novo* LoF (dnLoF) and *de novo* missense variants (dnMis) as compared to siblings, while the Social/Behavioral class displayed the lowest enrichment compared to siblings, though we found significant burden in all four of our classes (Figure 3B, left; FDR = 0.01/0.01/0.07/0.02). Meanwhile, rare inherited LoF and missense variants only display a statistically significant increase in the Mixed ASD with DD class, with a noticeably higher count per proband, suggesting a role of inherited SNPs in the outcomes of autistic children in this class (Figure 3B, right; FDR = 0.87/0.39/0.96/0.034). Our analyses differentiate the two classes with greater developmental delays, showing that the Broadly Impacted class has more high-impact *de novo* variants specifically, while the Mixed ASD with DD class has a combination of high-impact *de novo* and rare inherited variants, suggesting a unique inherited component for the children in this class.

We additionally observed differences across categories of LoF constraint, including potential significance masked by grouping the heterogeneous classes of probands together. For the purpose of this analysis, we leveraged the gene-level measure of probability of loss-of-function intolerance (pLI)^43^. We assigned genes with pLI >= 0.5 into one of two categories: high constraint genes (pLI >= 0.995), and intermediate constraint genes (0.5 <= pLI < 0.995). When we examined the burden of dnLoF variants across the classes in high constraint genes, we observed a pattern consistent with our findings above: Broadly Impacted displays the greatest burden counts in high constraint genes, though there is a significant increase relative to siblings among all autistic classes (Figure 3C, left; FDR = 0.009/0.007/0.01/0.007, OR = 3.64/6.40/2.76/3.70). This finding is consistent with prior work showing an excess burden of mutations in high constraint genes among probands^9,44^. However, previous work identified no significant increase in intermediate constraint genes in probands. By separating heterogeneous classes, we found that the Moderate Challenges class is significantly enriched for dnLoF variation in genes of intermediate constraint (Figure 3C, right; FDR = 0.048/0.26/0.12/0.18, OR = 2.8/1.9/2.25/1.99), suggesting that perhaps less essential genes are impacted in this class of individuals.

The genetic differences observed across classes support the importance of separation of individuals in order to identify coherent genetic architecture underlying clinical phenotypic presentation.

### Unique gene sets and cell biological processes are associated with phenotypic heterogeneity

Further insight into specific genes and processes dysregulated by the variants across the phenotype classes came from investigating the count burden of *de novo* and inherited variants in ASD-relevant gene sets^9,19,45^ (Supplementary Table 4). Though all four classes display an enrichment of *de novo* variation among ASD-related gene sets compared to non-autistic siblings, there are clear differences in the levels and patterns of enrichment between classes (Supplementary Table 5). For example, all autism-specific gene sets have significantly higher dnLoF mutation burdens in the classes with greater developmental delays, suggesting that cognitive outcomes are associated with rare high-impact mutations in a small subset of relevant genes (Figure 4A; Satterstrom: FDR = 0.05/0.05/0.004/0.004; ASD risk genes: FDR = 0.09/0.045/0.04/0.02). Odds ratio analysis further subdivides the classes, with ORs for dnLoF variants being greatest in the classes with greater developmental delays (Figure 4B, left; FDR = 0.18/0.003/0.02/0.002, OR = 4.86/27.14/9.46/18.9), while the ORs for *de novo* synonymous variants are uniformly distributed, with no significant increases over non-autistic siblings (Figure 4B, left; FDR = 0.78/0.78/0.78/0.78, OR = 0.75/1.65/1.37/1.45). Furthermore, *de novo* variants in the Fragile X Mental Retardation Protein (FMRP) target genes are strongly associated with the Broadly Impacted class, while inherited LoF variants in the same gene set are enriched in the Social/Behavioral class (Supplementary Fig. 6; Broadly Impacted: FDR = 0.014, Social/Behavioral: FDR = 0.029). Odds ratios further support the distinct strong dysregulation of FMRP target genes in the Broadly Impacted class, but more weakly in all the other classes, further pointing to the genetic differences between the two classes with the most developmental delays (Figure 4B, right; Broadly Impacted: FDR = 0.001, OR = 13.76; other: FDR = 0.003/0.003/0.004, OR = 6.15/6.37/6.28). Individuals with Fragile X syndrome exhibit developmental delays and intellectual disabilities as seen across both the Broadly Impacted and Mixed ASD with DD classes, but additionally tend to display mood disorders such as anxiety and impulsive, hyperactive, and aggressive behaviors^46^, which we observe only in the Broadly Impacted class. As expected, the non-autistic sibling group had no significant enrichments over probands across all tested gene sets.

**Figure 4.**
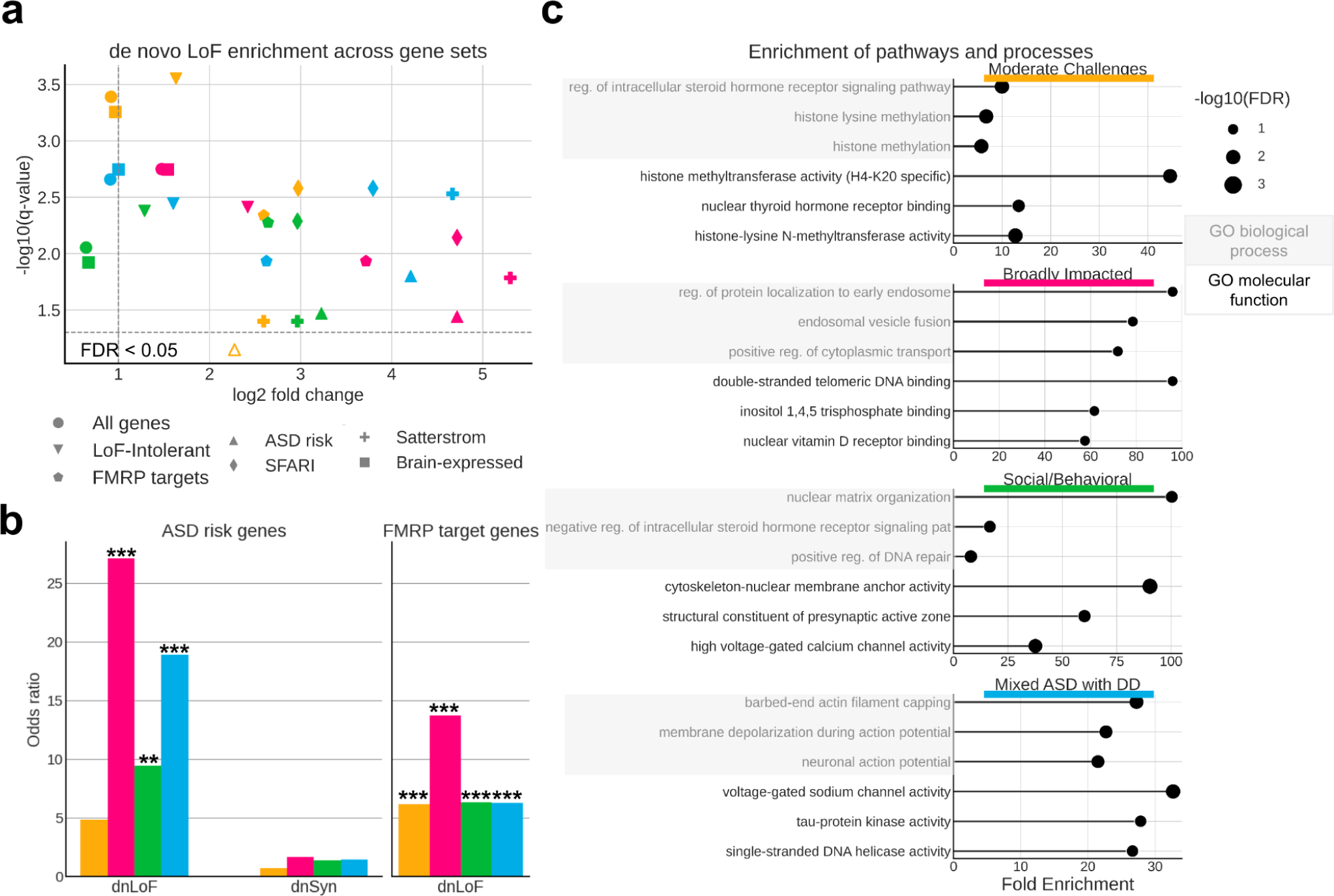
**a**, Scatter plot displaying enrichment versus significance of *de novo* LoF (dnLoF) burden for each class and gene set. We extracted 7 relevant gene sets and computed the aggregated dnLoF burden for each individual across every gene set. Log-transformed fold change (x-axis) and q-values (y-axis) were computed relatively to non-autistic siblings using a one-sided independent t-test. Gene sets in classes with FDR > 0.05 are shown below the dotted line (FDR = 0.05) and are indicated by open shapes. **b**, Odds ratios (y-axis) across classes for ASD risk genes (left) and FMRP target genes (right). We show similar trends in other autism-specific gene sets (Supplementary Fig. 5). Odds ratios for *de novo* synonymous (dnSyn) variation are also displayed for ASD risk genes. **c**, Top cell biological processes and molecular functions (GO) enriched for dnLoF in each autism class. Gene sets for GO enrichment analyses include all protein-coding genes impacted by high-confidence *de novo* loss of function or pathogenic missense variants present in individuals from each class. The plots display fold enrichment values (x-axis) and log-transformed FDR values (bubble size). Terms were selected by FDR and sorted by fold enrichment. For the Moderate Challenges, Social/Behavioral, and Mixed ASD with DD classes, an FDR cutoff of 0.05 was used, while a cutoff of 0.1 was used for the Broadly Impacted class due to its smaller sample size. Shaded boxes represent GO biological processes, and unshaded boxes represent GO molecular functions.

Molecular pathways affected by the distinct genetic variation observed across phenotypic classes also suggest different underlying biological mechanisms. Analysis of biological processes impacted by high-confidence *de novo* LoF or damaging missense variation in each class reveals little overlap in the top enriched biological processes and no overlap in top molecular functions between all four classes, suggesting that impacted genes represent pathways uniquely associated with class-specific phenotypes (Figure 4C; Supplementary Table 6). In particular, the Social/Behavioral class is highly enriched for processes of nuclear matrix and chromatin organization (FE = 100.4/3.9, FDR = 1.6e-02/2.2e-03), positive regulation of DNA repair (FE = 8, FDR = 1.0e-02), synapse-related molecular function (FE = 60.2, FDR = 1.5e-02), and core promoter sequence-specific DNA binding (FE = 16.7, FDR = 2.2e-03). The Moderate Challenges class displays strong enrichment for histone methylation (FE = 6.7, FDR = 2.6e-03) in addition to chromatin organization (FE = 3.4, FDR = 4.2e-4). In contrast, the Mixed ASD with DD class is defined by processes of neuronal action potential and membrane depolarization (FE = 21.5/22.7, FDR = 7.8e-03/7.7e-03), negative regulation of protein depolymerization (FE = 15.5, FDR = 2.0e-03), and voltage-gated sodium channel activity (FE = 32.7, FDR = 2.8e-03). Overall, our analysis directly suggests hypotheses for specific biological dysregulations underlying each autism class, providing a framework for directed examination of mechanistic insights in continuing autism research.

### Developmental gene expression patterns align with class-specific clinical milestones

We found that genes affected by variants in each ASD phenotypic class are associated with unique patterns of gene expression trajectories throughout brain development (Figure 5; Supplementary Table 7; Supplementary Fig. 9). This analysis leveraged cell-type-specific developmental gene expression trajectories of the human prefrontal cortex (Figure 5A, Methods)^47^. We found that the Mixed ASD with DD class is enriched for *de novo* LoF variants that affect genes expressed in all major prefrontal cortex cell types and mostly during the fetal and neonatal stages, with declining expression later in development (termed “Trans Down” and “Down” patterns in Herring *et al.*; Figure 5A bottom; Figure 5B). In contrast, the Social/Behavioral class is only enriched for LoF variants in genes highly expressed postnatally (termed “Trans Up” and “Up” genes; Figure 5A top; Figure 5B) in inhibitory interneurons (MGE). These developmental gene expression patterns align with the developmental clinical milestones of the classes: the Mixed ASD with DD class has the latest average age of developmental milestone attainment (Figure 1C) and, relatedly, the youngest average age of diagnosis (Figure 2B). In contrast, the Social/Behavioral class, which has variants in later expressed genes, shows less impact on early development, with later diagnosis ages and developmental milestones in line with non-autistic siblings. The Moderate Challenges class similarly displayed enrichment in mostly prenatal gene sets (“Down” and “Trans Down” genes), though the genes impacted within this class tend to be of lower evolutionary constraint than genes impacted in Mixed ASD with DD, which may contribute to the differences in outcomes (Moderate Challenges median pLI = 0.75, Mixed ASD with DD median pLI = 0.95; p-value = 0.026). Finally, Broadly Impacted displayed significance in both directions. The pattern of dysregulation includes most in-utero stages, aligning with the Mixed ASD with DD class, as well as postnatal stages, indicating broad dysregulation across developmental stages and cell types. Our findings thus demonstrate that there are class-specific differences in the developmental timing of genes that are dysregulated, which correspond to clinical milestone and outcome differences between the classes.

**Figure 5.**
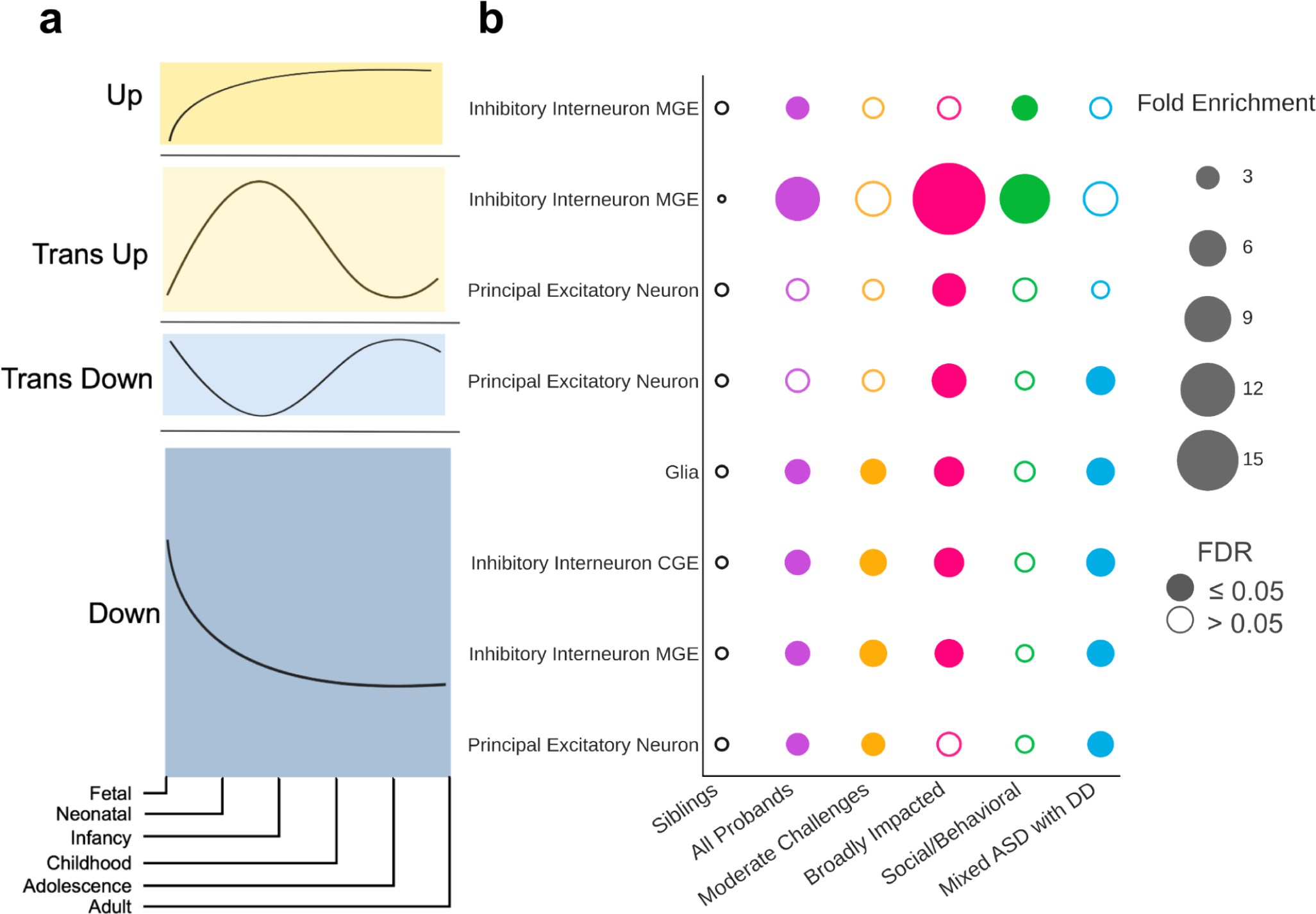
**a**, Trends from Herring *et al*. representing the gene expression trajectories of brain development genes differentially expressed across developmental stages. Gene expression trajectories follow one of four general patterns: “Up” (first), “Trans Up” (second), “Trans Down” (third), “Down” (fourth). Trends are measured across the 6 stages of development (x-axis): fetal, neonatal, infancy, childhood, adolescence, and adulthood. **b**, Patterns of *de novo* LoF variant enrichment across classes (x-axis), major cell types of the prefrontal cortex (y-axis), and gene expression trends (y-axis). For each class, we computed the fold enrichment (bubble size) and corrected p-values (FDR) of variant burden compared to non-autistic siblings. Open circles indicate FDR > 0.05 (not significant), and closed circles indicate significant enrichment (FDR ≤ 0.05). Each column is colored by the corresponding phenotypic class color, with purple representing the combined pool of all probands. Cell type and trend combinations with no significant enrichment in any class are not shown.

## Discussion

Unraveling the complexity of autism is a particularly challenging yet critical task for supporting the needs of autistic individuals and understanding the biology, inheritance, trajectory, and phenotypes of the many forms of the condition. A person-centered modeling approach powered the classification of over 5,000 autistic children from ages 4-18 based on a broad set of phenotypes that interact and co-occur, revealing distinct genetic and developmental signals tied to clinical presentations.

The classes we have defined, validated, and replicated reflect a robust quantitative analysis of broad phenotypic information on a large sample. The clear separation of genetic signals for each class establishes a concrete set of hypotheses that can be tested with larger cohorts and more comprehensive phenotyping. It is therefore crucial to both expand the cohort size and the quality and breadth of the phenotyping in order to more completely capture the full diversity of the autistic population. Coupled with genetic data, future studies will be better equipped to power significant associations between phenotype classes and genotypes, beyond single gene models. Extensive digital phenotypes and longitudinal data, combined with the power of large, genetically profiled cohorts will drive improved phenotypic and genetic resolution. Additional genetic information from whole genome sequencing offers broad potential for discovery of regulatory mechanisms contributing to phenotypic outcomes.

While all classes have some enrichment for common and rare variants, we find distinct signals which define each autism class. The Moderate Challenges class is uniquely enriched in common variants associated with autism and has the highest signal of rare variants in genes of lower evolutionary constraint. The Social/Behavioral class demonstrates the highest signal for common variants associated with co-occurring ADHD and depression, with modest enrichment for high-impact *de novo* and rare inherited variants, especially in neuronal genes predominantly expressed postnatally. In contrast, the Mixed ASD with DD class is highly enriched for all high-impact rare variation, both *de novo* and inherited, particularly in neuronal genes primarily expressed in utero. Finally, the Broadly Impacted class has the greatest enrichment for high-impact *de novo* variants, specifically in highly constrained genes and FMRP target genes. By leveraging a person-centered approach, we reveal that these robust, phenotypically distinct classes display characteristic patterns of genetic variation. Unlike previous approaches, our analyses directly associate genetic signals with sets of co-occurring phenotypes, and further implicate specific affected pathways, brain cell types, and developmental stages. We propose that clinical milestones can be partially attributed to the expression trajectories of brain development genes affected by deleterious variation in each autism class.

Our findings point to new directions for biologists and neuroscientists to pursue in order to gain insights into the underlying neurobiology of particular autism presentations, and offer the potential for more precise clinical diagnosis and guidance. Future research could also examine how interventions may differ between the classes. We have demonstrated that person-centered quantitative phenotypic analysis, combined with matched genetic sequencing data at scale, is crucial for uncovering phenotypic classes and their corresponding genetic factors that contribute to the broad heterogeneity of autism.

## Methods

### Participants

For the phenotype analyses, we restricted our sample to children aged 4 to 18 from the SPARK cohort^33^ which includes both autistic individuals and their non-autistic siblings. The SPARK Phenotype Dataset V9 battery assays core autistic traits (social, language-related, and repetitive behaviors), cognitive measures, and co-occurring behavioral features (such as anxiety, aggression, etc). Measures in SPARK with a sufficiently high overlap among participants were selected to maximize the cohort size while maintaining breadth of phenotype data. These measures included the Background history (Bhx, a form which asks parents to report on ages of developmental milestones), Social Communication Questionnaire-Lifetime (SCQ)^34^, Repetitive Behavior Scale (RBS-R)^35^, and Child Behavior Checklist 6-18 (CBCL)^36^. More details on the phenotyping measures are below. All autistic children with data from these four phenotype measurement tools were included in the mixture modeling analysis. The same sample of individuals was measured repeatedly. To maximize our sample and feature sizes, we excluded features with less than 90% completeness across the cohort, and only included individuals with complete measures across the remaining features. Our resulting cohort included 239 item-level and composite phenotype features and 5,392 individuals in SPARK alone (*n* = 4,174 males). The cohort had a mean age of 8.56 years with a standard deviation of 3.15 years. Our sibling cohort comprised 1745 paired siblings (related to at least one proband in the autistic cohort) with available WES data. Only the Bhx and SCQ were available for SPARK siblings out of the four phenotypic measures. For trio-based analyses, we only used complete trios in SPARK (*n* = 1,756 probands, *n* = 856 paired siblings). In both our phenotypic and genetic analyses, only individuals who passed QC and had complete data were included.

### Mixture modeling analyses

#### Phenotypes

We applied a finite mixture modeling analysis based on the phenotype features described above. The Background history form provided information on numerous developmental milestones including, but not limited to, age first walked, age first talked, age when combined phrases, age when bladder trained, etc. These features are numerical and measure parent-reported age (in months) when developmental milestones were achieved. The SCQ is a widely-used measure covering primarily social behavior difficulties and social communication, and repetitive/restrictive behaviors to a lesser extent. The questionnaire consists of 40 binary questions pertaining to social behaviors, communication, social interaction, sensory experiences, etc. The total score ranges from 0-39, with a higher score indicating higher impairment (more difficulties). We excluded the first question of the SCQ (q01_phrases), which is not used in the calculation of the total score. The RBS-R is a questionnaire covering six factors of behavior: self-injury, sameness, restrictive, repetitive, stereotyped, and ritualistic behaviors. It includes 43 categorical questions (rated 0-3) which load into six composite scores covering each area of behavior. The total score for the RBS-R ranges from 0-129, where higher scores indicate higher impairment (more difficulties). Finally, the CBCL 6-18 is a rating scale with 144 questions (121 of which were implemented in this study due to the exclusion of text-based questions) covering many co-occurring traits and conditions, including ADHD, OCD, depression, anxiety, somatic problems, etc. This questionnaire only covers individuals between 6 and 18 years of age; however, some individuals whose age is marked as less than 6 years old in our cohort had this measure available due to later administration of the test after recruitment. CBCL questions are measured on a scale from 0-2, where higher scores indicate more frequent behavior. These are summed up into 20 standardized composite t-scores each ranging from 0-100.

#### Mixture model and covariates

Features across the four phenotype measures were retrieved and integrated for each individual. The resulting matrix covered 5,392 individuals and 241 features (239 training features, 2 covariates). We used the StepMix package (version 1.2.5)^48^ to fit a model and predict labels for each individual using a General Finite Mixture Model (GFMM) approach, which is a specialized case of a generative model. This model expands upon the Gaussian Mixture Model by allowing different probability density functions to model continuous features (Bhx, RBS-R, CBCL), binary features (SCQ), and categorical features (RBS-R, CBCL). The final model separates features into data types, and models each feature with an appropriate probability density function (gaussian, binomial, or multinomial). The GFMM also has light statistical assumptions and is inherently a person-centered approach. We trained a 1-step StepMix model with covariates (sex and age at evaluation in years) and the following hyperparameters: n_init = 200 (200 models with different random seeds are trained and the optimal model is selected), and n_components = 4 (the number of class distributions, a hyperparameter which was tuned and selected through a multi-step process detailed below). The model predicts four probabilities for each individual (one for each latent class), and takes the max over these values to assign each individual to a single latent class.

#### Exploratory class enumeration

The main tunable hyperparameter of the GFMM is the number of latent classes. We tune this hyperparameter in a thorough, multi-step process which considers various statistical measures of model fit and indicators of optimality. We evaluated models with 1-12 components and computed the validation Log Likelihood (LL), Alkaline information criterion (AIC), Consistent AIC (CAIC), Bayesian information criterion (BIC), Sample-Size-Adjusted BIC (SABIC), Likelihood Ratio Test statistics (LMR-LRT), and more (Supplementary Table 1, Supplementary Fig. 1) across these models, averaged over 50-200 independent runs with randomly generated seeds.

The LL was computed over 200 independent runs for each model using a cross-validation scheme with 3 folds, where a model was fit on ⅔ of the cohort data, and tested on the remaining ⅓ of the data. The log likelihood was then computed and summed across individuals in each class.

The AIC, CAIC, BIC, and SABIC were computed over 200 independent runs using the log likelihood, but with added terms which penalize complexity (number of parameters) to different extents, which, in the case of the BIC/SABIC, is scaled in accordance with the sample size. The LL, AIC, CAIC, BIC, and CABIC (Supplementary Fig. 1) can be evaluated according to the elbow criterion: we looked for the point where the marginal gains in fit are diminished, causing an elbow in the plot. This point is located around 4-6 components in all five plots.

The Lo-Mendell-Rubin Likelihood Ratio Test (LMR-LRT) was performed over 50 independent runs of the model with randomly generated seeds. A Chi Square test statistic was computed for each pair of consecutive models (e.g. n_component=2 and n_component=3). The resulting p-value indicates whether the latter, more complex model offers a better fit over the simpler model, suggesting guidance for selection of the number of components in the model. The LMR-LRT results were evaluated according to the point where the average p-value increases above the cutoff for significance (alpha), marking the model with the best balance of fit and complexity. For our data, this point clearly occurred at n_components = 4, suggesting compromised fit for the 5 and 6 component models as compared to the 4 component model.

We additionally measured average posterior probability (AvePP) and entropy across candidate models. The AvePP was computed by extracting the maximum posterior probability per individual, and averaging the probabilities for the pool of individuals in each class. A higher posterior probability indicates higher confidence of class assignment. The entropy per model was also computed, since higher entropy indicates better fit. However, neither AvePP nor entropy were used as measures for model selection, but rather as confirmation of model fit. We also recorded the count and proportion of the smallest class across all models. A model with a higher number of components tends towards fragmentation of class sizes, so we sought to balance meaningful class sizes with adequate phenotypic decomposition.

#### Interpretation

Finally, a strategy that is additionally valuable involves rendering an interpretation of each class in each candidate model, and evaluating the utility, clarity, and validity of the resulting model interpretations. We consulted with clinical collaborators on the interpretability of multiple candidate models, and the four class solution offered the best phenotypic separation and most clinically coherent classes. Less complex models offered insufficient phenotypic reduction, while more complex models overfit our data and resulted in fragmented classes which were phenotypically fine-grained. The final four class breakdown resulted in the following class sizes: class 0: n = 1976 (37%); class 1: n = 554 (10%); class 2: n = 1860 (34%); class 3: n = 1002 (19%) (Supplementary Fig. 7). Sex breakdown and percentages by class is as follows: class 0: 1459 males (78%), 401 females (22%); class 1: 441 males (80%), 113 females (20%); class 2: 1475 males (75%), 501 females (25%); class 3: 799 males (80%), 203 females (20%). Age and sex breakdown by class can be found in Supplementary Fig. 8.

As we outline above, there is not one clear indicator for this parameter choice. We evaluated a rigorous combination of statistical measures, combined with interpretability, to select our final model: a four component model which provides the best balance between model fit, complexity, and interpretation.

#### Phenotype categories

In order to phenotypically define and interpret our model outputs, we grouped the 239 features used in training the GFMM into 7 factors previously defined in literature^35,37–39^. Our final categories included: limited social/communication, restricted/repetitive behavior, attention, disruptive behavior, anxiety/mood, developmental delay, and self-injury. Each category is a composite of features assigned through prior grouping evidence in previous works. Some categories more clearly correspond to a specific phenotyping test: for example, disruptive behavior included questions drawn from the CBCL, and developmental delay is primarily the milestones from the Background history form, while other categories included a mix of measures: e.g. limited social/communication is a hybrid SCQ/CBCL category.

To more thoroughly examine how the factors define the four phenotype classes, we performed the following analysis: first, we computed the enrichment (upper tail significance) or depletion (lower tail significance) of each feature in each class relative to the other three classes. For binary variables, a one-sided binomial test was used. For categorical and continuous variables, a one-sided independent t-test was used. Multiple hypothesis correction was performed for each autism class and direction of enrichment with a Benjamini-Hochberg correction. We then computed the proportion of features with upper-tail and lower-tail significance in each category and class. At this stage, features were excluded based on 3 criteria: (1) features which had no significant enrichment or depletion in any class, (2) continuous or categorical features with Cohen’s d values between −0.2 and 0.2 across all classes, and (3) binary features with fold enrichment of less than 1.5 across all classes. This resulted in 220 contributory features which were used in subsequent analyses. After computing the proportion of features enriched or depleted in each class and category, we negated the depleted proportions and summed them with the positive enriched proportions. This enabled us to compute a total proportion and direction of significant features for each category and class. This score represents the “affinity” of the class towards a phenotype category, relative to other autistic children. This analysis allowed us to summarize the phenotypic trends which define each class, and render interpretations of the model outputs, including a thorough characterization of core, associated, and co-occurring phenotypes.

#### Naming

We consulted with clinicians, parents of autistic children, autistic adults, and other researchers to determine naming that would be accurate, descriptive, and non-ableist.

#### Phenotypic Replication on the Simons Simplex Collection

We replicated the four SPARK phenotype classes in an independent cohort – the Simons Simplex Collection^40^. We first extracted, processed, and integrated the same phenotype measures from SSC that were used in the SPARK model: SCQ, RBS-R, CBCL 6-18 (only composite scores), and a form consisting of parent-reported timing of developmental milestone attainment. We manually processed the developmental milestones form, which initially included open responses and inconsistent time scales across features. We identified 108 phenotype features which were present in both SPARK and SSC. To demonstrate the generalizability of the SPARK model to SSC, we trained a model only on the SPARK data for the 108 common features with the same hyperparameters as the model described above (*n* = 6,393 individuals in SPARK with complete data for the 108 features). We compiled the same features for *n* = 861 individuals in SSC who had complete data across the training features. We then applied the trained model to the SSC test data, and predicted a class label for each individual from SSC. At this point, we excluded non-contributory features post-hoc based only on the training data and according to the same inclusion criteria described above. We obtained the proportion and direction of enrichment for each class and category across both SPARK and SSC. The Pearson’s correlation coefficients between SPARK and SSC for each of 7 phenotype categories are 0.98, 0.92, 0.94, 0.92, 0.89, 0.97, 0.98. Next, we obtained a significance of overall model similarity with a permutation test: we randomly shuffled the class labels of SSC participants for n = 10,000 repetitions, obtaining a distribution of chance correlations between SPARK and SSC. We then compared the true correlation between SPARK and SSC (unshuffled labels) to the chance correlation distribution, allowing us to obtain a p-value for overall model similarity which accounts for chance correlations.

### Phenotypic validations

#### External validation with Basic Medical Screening

Phenotype measures which were available for the majority of our cohort but were not included in the training or construction of the classes were used for external validation of the classes, as well as for exploratory phenotypic associations of clinical and diagnostic variables. One phenotyping form available in SPARK is the basic medical screening (BMS). This questionnaire includes binary (yes-or-no) indicators of diagnoses ranging across medical, morphological, cognitive, motor, mental health, neurodevelopmental conditions, sleep and eating disorders, among others. Of our cohort sample, the majority of individuals (*n* = 5,381) had a complete or nearly complete BMS form available. To explore the associations of clinical diagnoses with the four classes, we computed the fold enrichment (FE) and p-values (one-sided binomial test) of each diagnosis within each class. To compute FDR values, we performed multiple hypothesis correction using Benjamini-Hochberg correction. We grouped select diagnoses of interest into three categories: neurodevelopmental, mental health, and co-occurring conditions.

#### External validation with additional self-report data

In SPARK, each enrolled individual provides a self- (or parent-) reported registration form upon enrollment, with baseline information covering a variety of phenotypic domains. Information from this form was not used in the training of our model or in the construction of our classes. We utilized this data to validate and explore phenotypic differences and similarities between the classes. In particular, we examined three variables of interest: diagnosis age (in months), cognitive impairment at enrollment (binary), and language level at enrollment (categorical). We computed the mean and confidence interval for each feature and class (Figure 2b) in order to characterize the differences between individuals as documented by their provided information at registration.

#### SCQ and Developmental milestones

Unlike most phenotype measures, the SCQ and Background history form were available for most non-autistic siblings, allowing for a baseline to compare to our latent classes. We compared the distributions of SCQ total scores (0-39) and ages (in months) when major developmental milestones were achieved – age when first walked and age when first used words. We performed hypothesis testing (one-sided independent t-test) with multiple hypothesis correction for each phenotype variable using Benjamini-Hochberg correction (Supplementary Table 2).

### Genetic analyses

#### Polygenic scores

The most recent, well-powered GWAS for ASD and several ASD-related traits and co-occurring conditions were used to compute polygenic scores for the subset of our cohort with genotyping array data and European ancestry (determined by a self-reported race of white) passing some basic quality control metrics (*n* = 2,293 autistic individuals, *n =* 3,179 non-autistic siblings). The set of siblings was chosen as all siblings below the age of 18 in the same family as the proband set, with available genotyping data. Genotypes were determined from Infinium Global Screen Array BeadChips available from SPARK^33^. Summary statistics were downloaded and uniformly processed before computing PRS with PLINK’s clumping algorithm^49^. Results were regressed by sex and the first 6 principal components to control for ancestry. All GWAS statistics used were based on populations of European ancestry. The full list of GWAS studies examined in this work can be found in Supplementary Table 3.

#### De novo variant calling and filtering

*De novo* variants (DNVs) were called using the HAT^42^ software for all complete trios in our proband and sibling cohort with whole exome sequence (WES) data (*n* = 1,680 probands, *n* = 801 paired siblings) from the Variant Call Format (VCF) files released by the SPARK Consortium. Briefly, this pipeline, optimized for whole genome sequencing (WGS) and WES data, uses variant calls from both DeepVariant^50^ and GATK HaplotypeCaller^51^ to identify variants which appear in both variant call sets in the child (genotype of 0/1 or 1/1), but not in either parent (both genotypes 0/0). Variants are then filtered based on several quality metrics including read depth, genotype quality score, and genomic regions. Variants in recent repeats, low complexity regions, and centromeres were filtered out. We used the default values for the filtering parameters of minimum depth = 10 and minimum genotype quality score (GQ) = 20. This pipeline identified an average of 2.71 DNVs per proband (std dev = 2.20) and 2.47 DNVs per sibling (std dev = 1.63). For filtering and quality control of DNVs, individuals with a high variant count (>3SD above the mean) were excluded from the analysis, and non-singleton DNVs (variants appearing in multiple SPARK families) were removed from further processing and analysis.

#### Rare inherited variation

The DNV pipeline was adapted to identify inherited variants using the same quality metrics. Inherited variants were identified as variants appearing in a child (genotype of 0/1 or 1/1) and in at least one parent (either genotype 0/1 or 1/1). They were then filtered in the same way as the DNVs to identify a high confidence set of inherited variants. We identified an average of 41,725.2 inherited variants per proband (std dev = 2,580.7) and 41,778.9 inherited variants per sibling (std dev = 2,314.6). In all analyses, rare inherited variants are defined as inherited variants with an allele frequency < 1% in gnomAD (v4.1.0).

#### Gene sets and resources

We looked for variant enrichment in a total of 7 different gene sets, including the set of all protein coding genes from Gencode v29 (Supplementary Table 4). The SFARI gene set was extracted from SFARIGene and includes category 1 genes only^19,22^. The Satterstrom gene set was retrieved from Satterstrom *et al.*^9^. The rest of the gene sets were retrieved from Werling *et al.*^45^, and include genes with pLI > 0.9 from ExAC^52^, predicted ASD risk genes (FDR < 0.3) from Sanders *et al*.^6^, and target genes of the fragile X mental retardation protein^53^. Finally, brain expressed genes were selected using the expression table from GTEx v.7 (gene median transcripts per million (TPM) per tissue)^54^. The genes selected were those whose expression in brain tissue was at least five times higher than the median expression across all tissues. Finally, the high and moderate constraint gene sets were selected using pLI scores (probability of being loss-of-function intolerant) from gnomAD^43^.

#### Count-based burden analyses

For analyses of *de novo* and inherited variation, we analyzed the distributions of variant count burden for each class of probands or siblings. After extracting *de novo* and inherited variants for each individual, we ran the variants through Ensembl’s Variant Effect Prediction (VEP)^55^ tool (Release 111.0) with the --pick flag (to obtain the most severe consequence) and the LOFTEE^43^ (v.1.0.4) and AlphaMissense^56^ plugins. To predict loss-of-function (LoF) variants, we flagged variants with the following predicted consequences: stop gained, frameshift variant, splice acceptor variant, splice donor variant, start lost, stop lost, and transcript ablation. We further subset the variants to only include those flagged as high confidence (“HC”) by LOFTEE. To predict high confidence missense mutations, we flagged variants with the following predicted consequences: missense variant, inframe deletion, inframe insertion, and protein altering variant. We further subset the variants to those predicted as “likely pathogenic” by AlphaMissense.

Some individuals had zero exome-wide calls for *de novo* variants (*n* = 76 probands and *n* = 55 siblings). In those cases, we accounted for their variant counts post-hoc (after variant effect prediction) in each count-based analysis of DNVs. Specifically, in each analysis we added a count of 0 for each proband or sibling with no DNVs to their respective class distribution prior to computing enrichments and hypothesis testing.

We aggregated the counts of *de novo* loss-of-function (dnLoF), de novo missense (dnMis), rare inherited LoF variants (inhLoF), and rare inherited missense variants (inhMis) for each individual. Modeling the variant counts as a distribution for each class and siblings, we performed hypothesis testing (one-tailed independent t-test) followed by multiple hypothesis correction for each variant class (Benjamini-Hochberg correction) to determine whether the variant count distributions significantly differed. To summarize the burden of high-impact mutations by class, we separately aggregated *de novo* variants (dnLoF and dnMis) and rare inherited variants (inhLoF and inhMis) for each individual, and computed the mean and standard error of burden per offspring for each class.

#### Gene set analyses

For analyses of specific gene sets, we first subset *de novo* and inherited variants to only include variants located in each set of genes. We then computed the gene-set-specific burden counts and repeated the hypothesis testing procedure to determine class enrichment for high-impact variants in each gene set. To summarize results across all gene sets and classes, scatter plots were constructed by computing the log-transformed fold enrichment (FE) of the means (each class compared to siblings) as well as log-transformed q-values from hypothesis testing (one-sided independent t-test) followed by multiple hypothesis correction for each gene set using Benjamini-Hochberg correction. All proband variant count distributions were tested against the sibling variant count distribution. Results and statistics for all classes and gene sets can be found in Supplementary Table 5.

#### Odds ratios

Odds ratios were computed for each phenotype class and gene set. Briefly, for each class and gene set we extracted the number of individuals in the case (proband class) with a variant in any of the genes from the gene set, as well as the count of probands with variants absent in those genes. We also extracted the present and absent counts for the control (non-autistic siblings). All comparisons were made for each class against siblings. P-values were corrected using Benjamini-Hochberg correction for each gene set and variant type (LoF, Syn) separately.

#### GO term analysis

To extract the top disrupted biological processes for each class, we flagged dnLoF and dnMis variants present in all protein-coding genes across individuals in each class. Taking the set of impacted genes (genes containing high-impact variants) for each autism class, we tested the hypothesis that class-specific gene subsets represent distinct pathways and biological processes. To extract the top GO biological processes and molecular functions for each class, we used ShinyGO 0.80^57^, which computes the fold enrichment and FDR enrichment values using a hypergeometric test, with all protein-coding genes as background. We selected the terms by FDR enrichment and sorted for top terms by fold enrichment values. We then selected the top 20 biological process terms and the top 20 molecular function terms for each class (Supplementary Table 6). For classes 0, 2, and 3 we used an FDR cutoff of 0.05. For class 1, we used an FDR cutoff of 0.1 due to the smaller class size. All results were generated using ShinyGO 0.80 on June 3, 2024.

#### Developmental gene expression analysis

We leveraged a single-cell human prefrontal cortex (PFC) dataset collected from postmortem tissues at 6 different stages of development^47^. We used Table S3 from Herring *et al*. to retrieve brain development gene sets (termed devDEGs) for PFC cell types which follow one of four general gene trend patterns defined by the authors: “Up” (increasing expression throughout stages, with highest expression in late stages), “Trans Up” (increasing expression in earlier stages, peak in middle stages, followed by a decline in later stages), “Down” (declining expression throughout stages, with highest expression in fetal stage), and “Trans Down” (declining expression in early stages, lowest expression in middle stages, and increasing expression in late stages). devDEGs were identified through a differential expression analysis between the 6 developmental stages (fetal, neonatal, infancy, childhood, adolescence, adulthood). Although 14 major trajectories were identified, we opted to use the four general gene trends for clarity and interpretability. We further combined clusters of cells into the four identified major cell type categories: principal excitatory neurons, inhibitory interneurons (MGE), inhibitory interneurons (CGE), and glia. This was done by taking the union of devDEGs associated with each major cell-type category. This simplification further improved interpretability of the results, allowing us to identify subsets of devDEGs disrupted in different PFC major cell types and gene expression trends. We conducted a count burden analysis of *de novo* LoF mutations in each phenotype class compared to non-autistic siblings, and computed fold enrichments and p-values (one-sided independent t-test). To compute FDR values, we conducted multiple hypothesis correction for each gene set using Benjamini-Hochberg correction. We additionally computed statistics for the pool of all probands compared to siblings. Full results (for both significant and insignificant celltype and trend combinations) can be found in Supplementary Figure 9. Fold enrichments, p-values, and q-values across all classes, cell types, and trends can be found in Supplementary Table 7.

To compare the constraint of genes impacted in the Moderate Challenges and Mixed ASD with DD classes, we extracted the genes with high-confidence dnLoF variants in each class for the following combinations of gene expression trends and cell types: down in principal excitatory neurons, down in inhibitory interneuron (MGE), down in inhibitory interneuron (CGE), and down in glia. These categories displayed enrichment for dnLoF variants across both classes. We extracted the pLI values for impacted genes in each class, and conducted a one-tailed Mood’s median test to test whether the difference in the distributions of pLI scores is significantly different between the two classes.

## Ethics

We received approval to access and analyze de-identified genetic and phenotypic data from the two cohorts from SFARI Base and the Princeton University IRB Committee in the Office of Research Integrity.

## Acknowledgements

We are grateful to all the families in SPARK, the SPARK clinical sites and SPARK staff. We appreciate obtaining access to the SPARK phenotypic and genetic datasets on SFARI Base. Approved researchers can obtain the SPARK population data set described in this study by applying at https://base.sfari.org. We are grateful for funding from the NIH NIGMS (R01GM071966), NHGRI (R01HG005998), Simons Foundation grant (395506), and the NIH NHGRI training grant T32HG003284.

## Data Availability

In order to abide by the informed consents that individuals with autism and their family members signed when agreeing to participate in a SFARI cohort (SSC and SPARK), researchers must be approved by SFARI Base (https://base.sfari.org).

## Code Availability

Analysis code and scripts used for this manuscript will be made available upon publication.

## Notes

### Competing Interest Statement

The authors have declared no competing interest.

### Author Declarations

The Office of Research Integrity of Princeton University gave ethical approval for this work.

