## Supplementary Figures for "Decomposition of phenotypic heterogeneity in autism reveals distinct and coherent genetic programs"

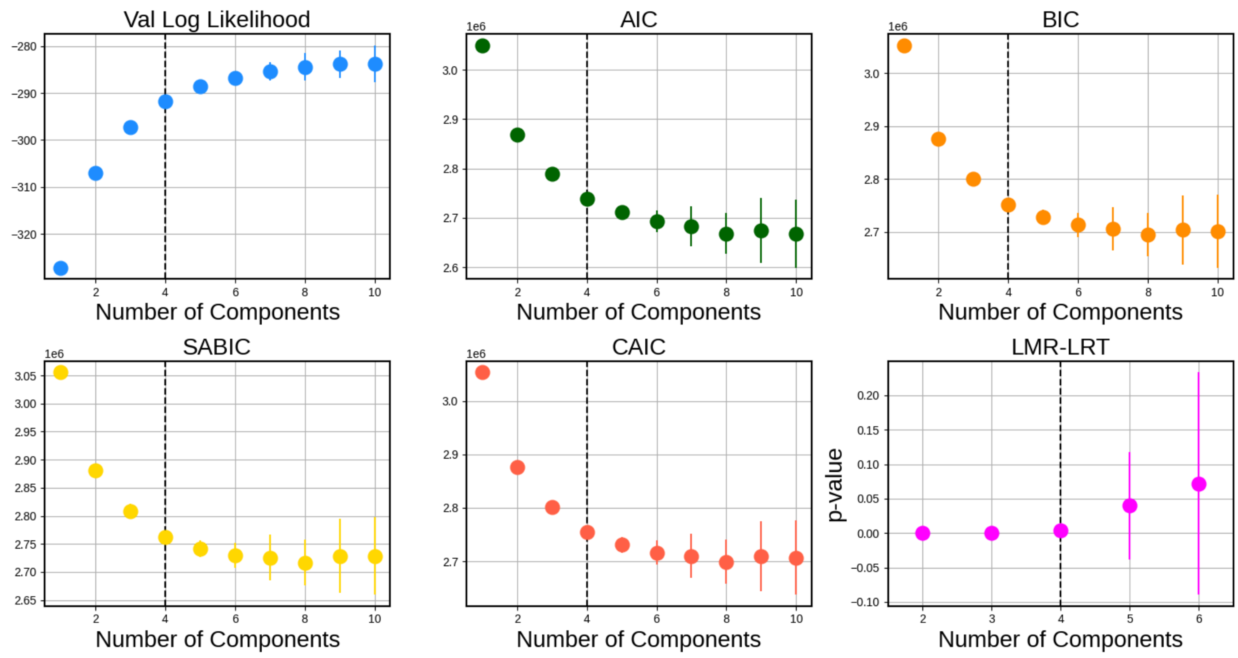


**<u>Supplementary Figure 1</u>: Exploratory class enumeration analyses suggest 4 components is a suitable parameter choice for the SPARK generative mixture model.** A crucial step in mixture modeling is tuning the number of model components through evaluation of fit metrics. Models were trained with varying numbers of components (x-axis) over 50-200 iterations with randomly generated seeds. The distributions are plotted (mean and standard error) for a variety of important indicators: the Validation Log Likelihood (LL), the Akaike Information Criterion (AIC), the Bayesian Information Criterion (BIC), the Sample-Size-Adjusted BIC (SABIC), the Consistent AIC (CAIC), and the Likelihood Ratio Test (LRT). The evaluation of these indicators combined with interpretation of each candidate model and class resulted in the selection of a 4 component model (shown as vertical dotted line in all plots).

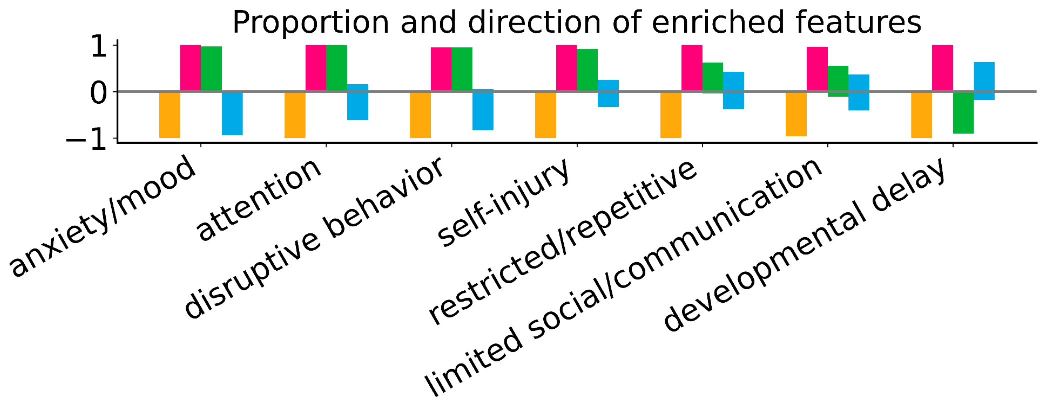

**<u>Supplementary Figure 2:</u> Variability of feature enrichment and direction.** To demonstrate variability in feature significance within phenotype categories and classes, enrichments and depletions are plotted for each combination of phenotype category and class, representing the proportion and direction of enrichment of features assigned to the category and computed within the class.

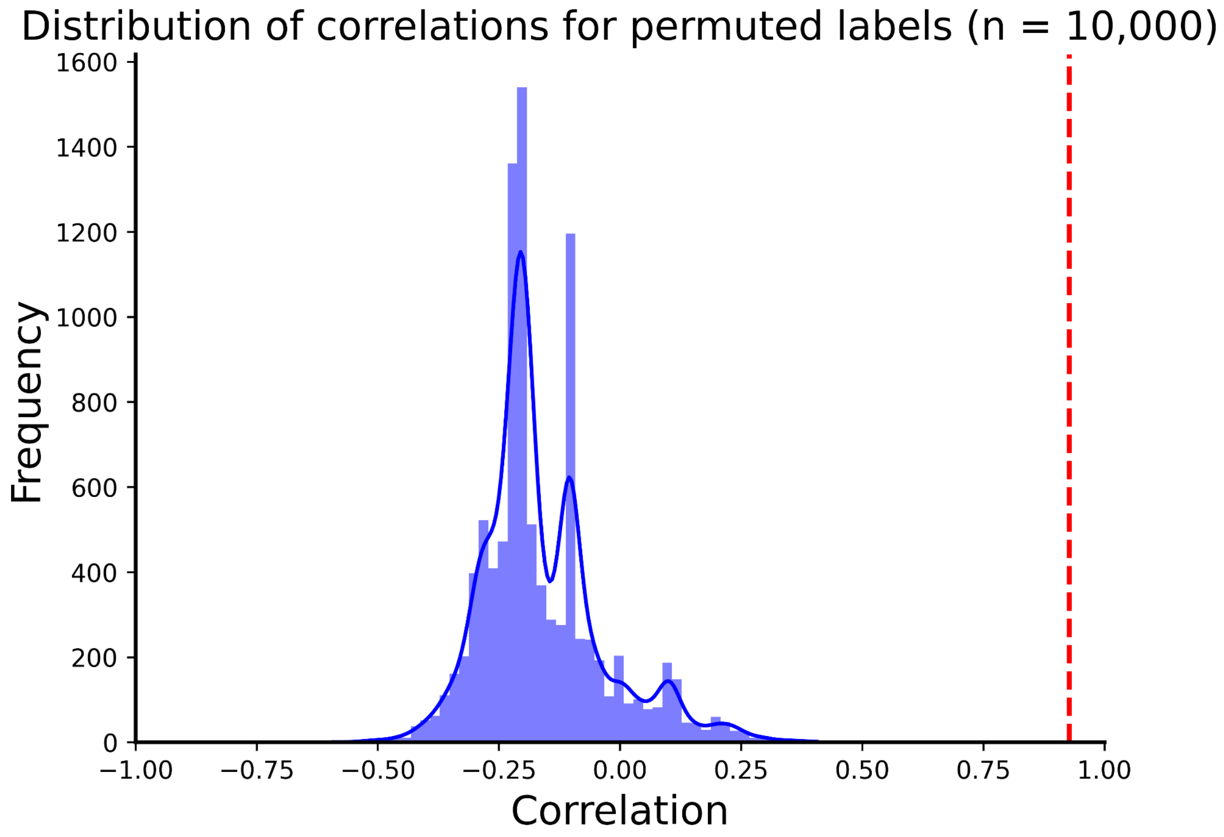


**<u>Supplementary Figure 3: Permutation analysis for SSC replication.</u>** Correlations between the feature enrichment of latent classes in SSC and SPARK were computed for 10,000 random permutations of the class labels in SSC. The dashed red line represents the correlation from applying the model trained on SPARK data to predict labels on individuals in SSC (r = 0.927), and yields a p-value < 1E-4 when compared to the chance correlation distribution.

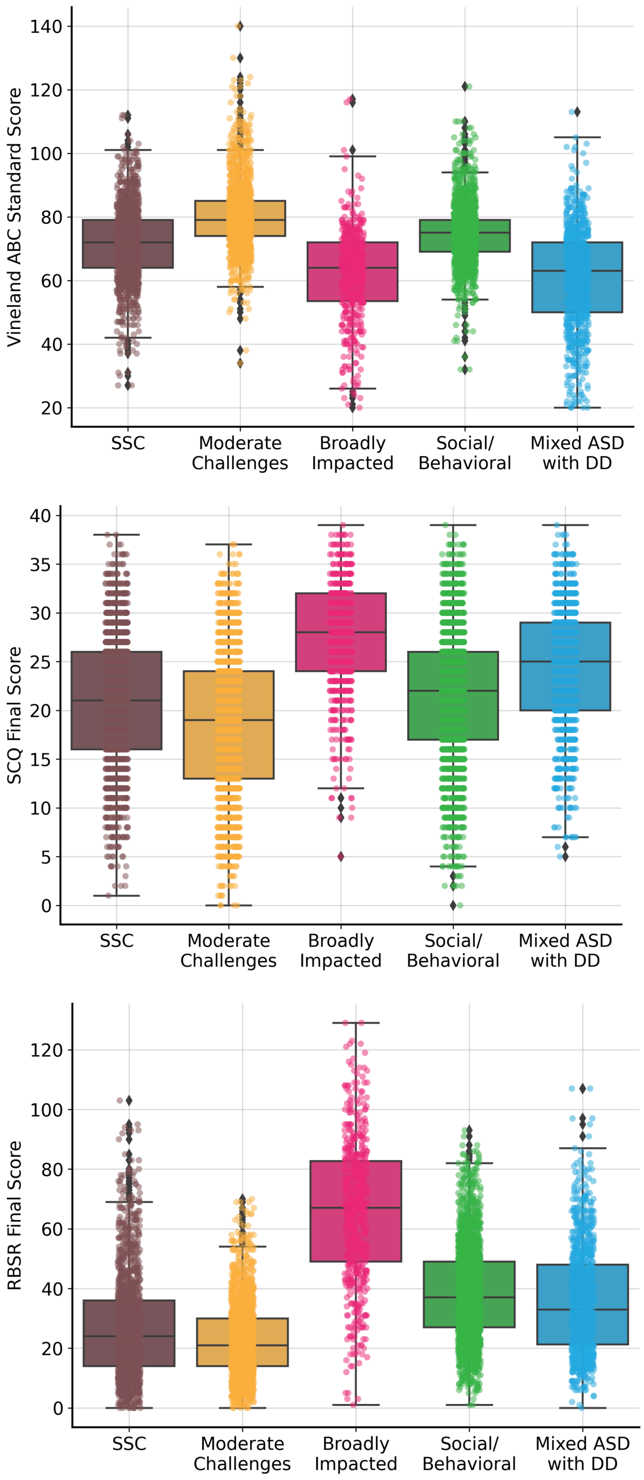


**<u>Supplementary Figure 4: SSC cohort vs SPARK classes.</u>** The ASD GWAS was trained in part on the SSC cohort, a group of autistic children most phenotypically similar to the Limited Impairment and Social/RRB classes of SPARK. The GWAS results are therefore likely to best reflect probands with similar phenotypes to SSC, which could explain why these classes have the highest autism PGS.

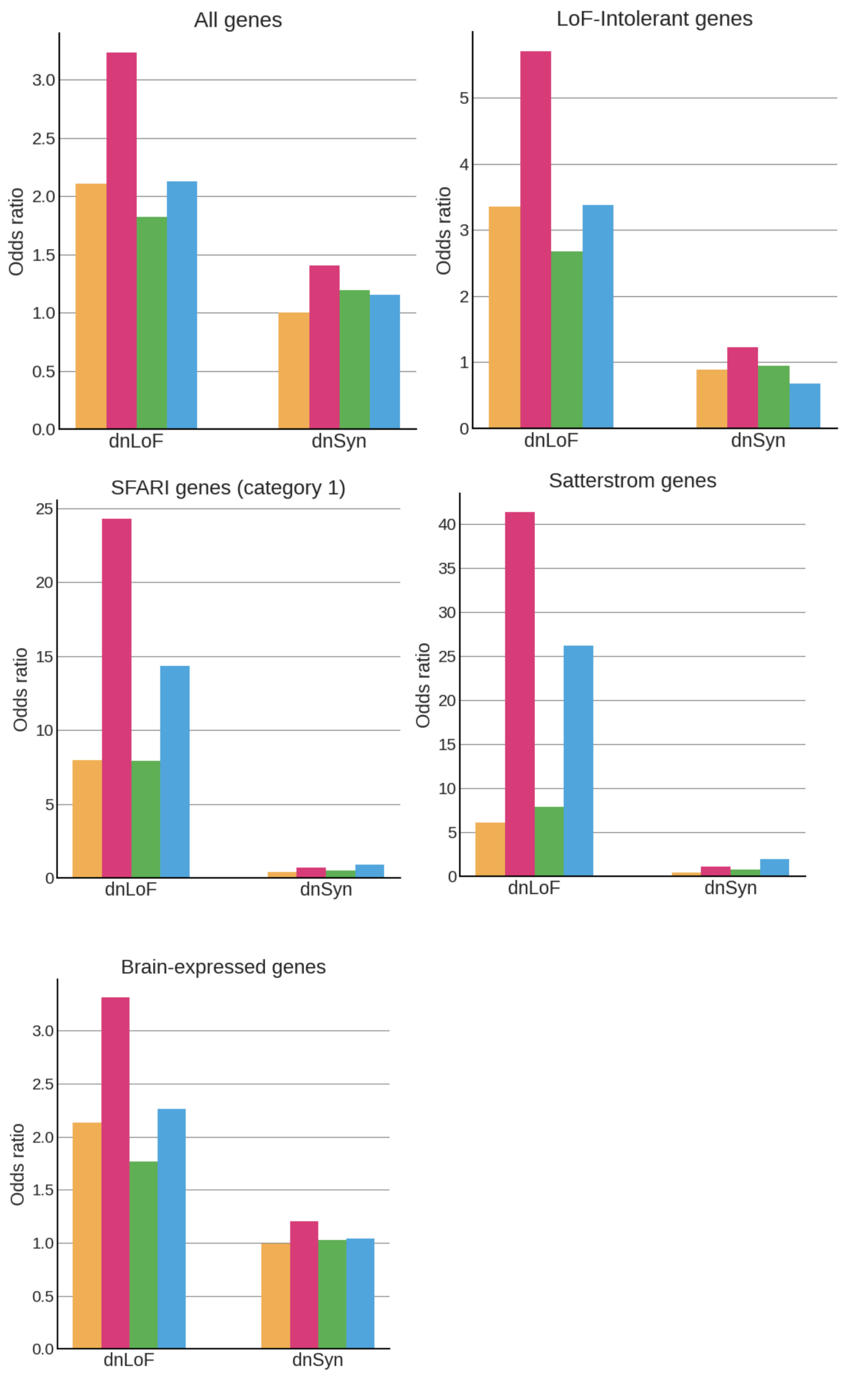


**Supplementary Figure 5: Odds ratios for all other autism-relevant gene sets included in this manuscript.** Odds ratios were computed for dnLoF and dnSyn variants in each autism class as compared to non-autistic siblings, across seven autism-related gene sets (five shown here, two shown in main Figure 4).

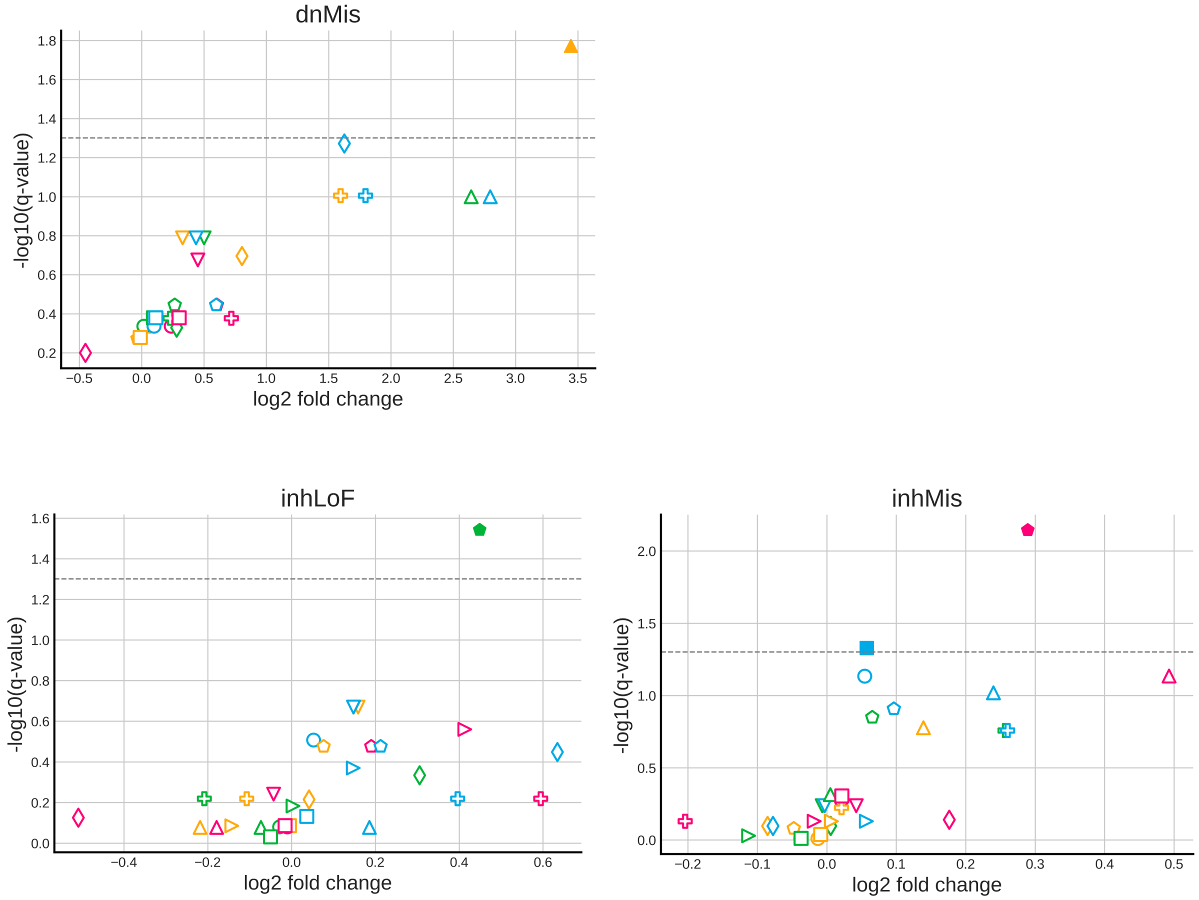


**<u>Supplementary Figure 6: Scatter plots for different types of variant burden within relevant gene sets</u>**. Scatter plots displaying enrichment versus significance of *de novo* missense (dnMis) burden, rare inherited LoF (inhLoF) burden, and rare inherited missense (inhMis) burden across classes and seven autism-relevant gene sets. We computed the aggregated burdens for each individual across every gene set. P-values and log2 fold change were computed relatively to non-autistic siblings using a one-sided independent t-test. Benjamini-Hochberg multiple hypothesis correction was then applied to compute the log-transformed q-values (y-axis). Empty shapes represent tests that did not pass multiple hypothesis testing with a threshold of 0.05.

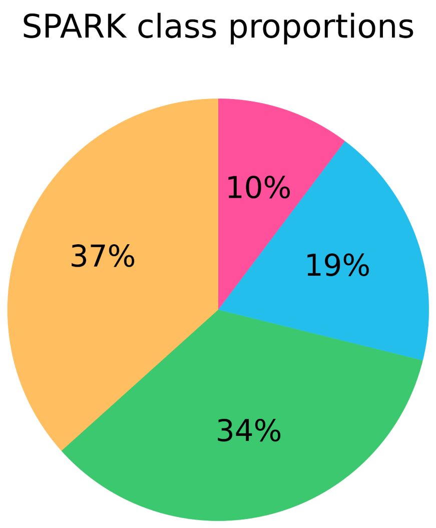


**Supplementary Figure 7: Pie chart of autism class percentages**. Breakdown of proportion of SPARK cohort individuals (total $n = 5{,}392$) assigned by the generative mixture model to each latent class. Classes are represented by the corresponding class colors, and numbers are rounded to the nearest percent.

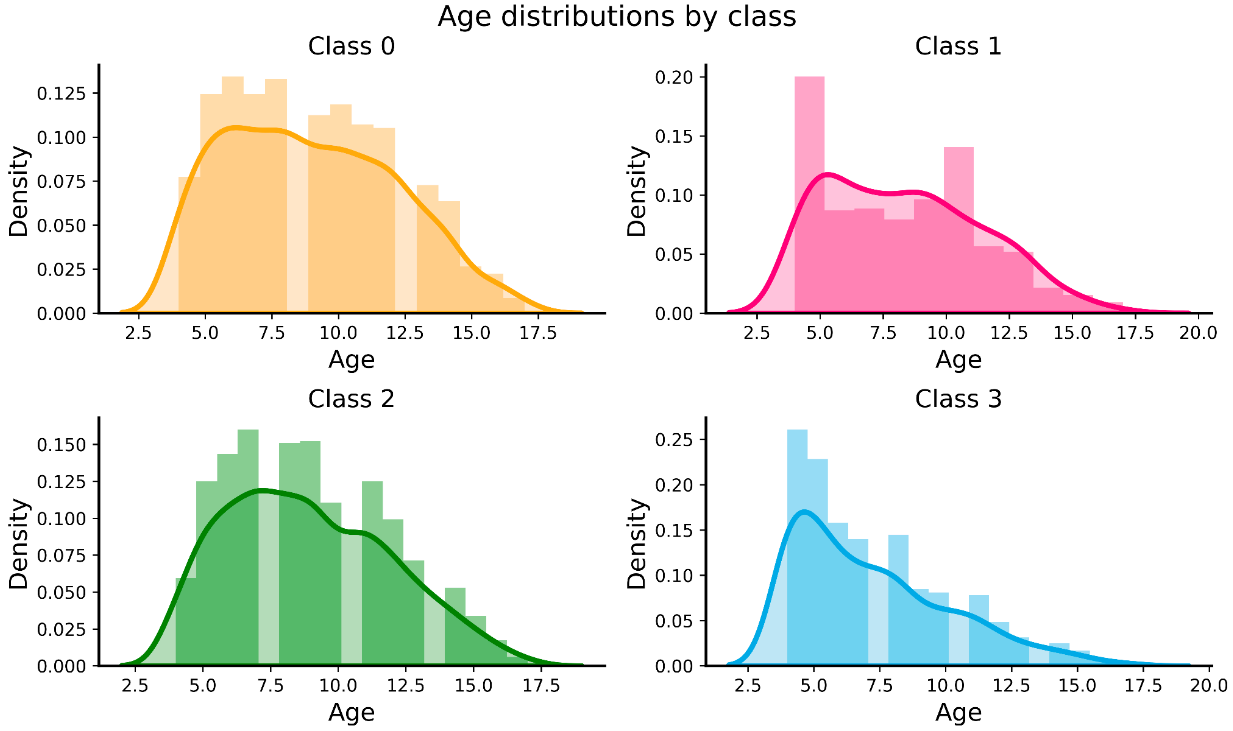

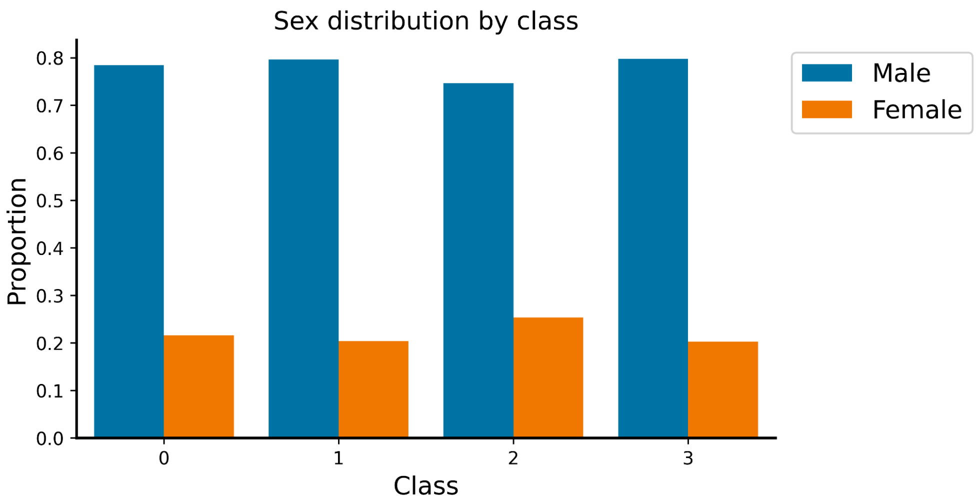


**Supplementary Figure 8: Age and sex distribution breakdown by class**. **a**, Density of age distribution for individuals assigned to each latent class. Colors correspond to class colors. **b**, Proportion of males and females for individuals assigned to each latent class.

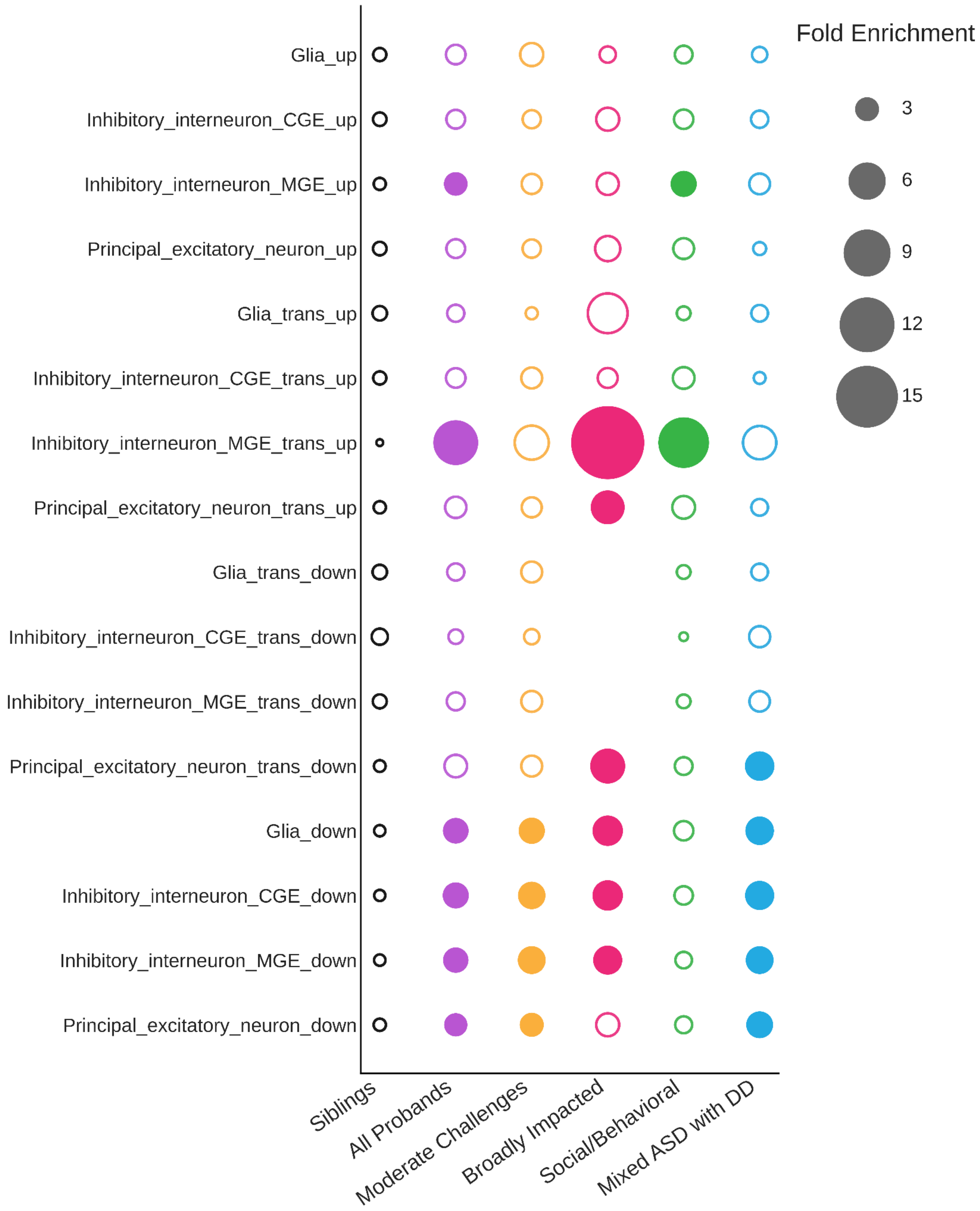


**<u>Supplementary Figure 9: Developmental gene trends and dnLoF enrichments across all measured PFC celltypes, gene trends, and phenotype classes.</u>** No circle represents lack of data. Open circle represents lack of significance (FDR > 0.05), whereas closed circle represents FDR <= 0.05. Colors correspond to class theme colors.

**Supplementary tables in separate files.**
